# Deciphering Environmental Health Factors Behind Unknown Etiology of Chronic Kidney Disease in South Asia: Plans for Epidemiologic Study

**DOI:** 10.64898/2026.03.28.26349626

**Authors:** Anirudh Mazumder, Sebastian Pintea, Aneesh Mazumder, Luke Chen, Jeffrey B. Kopp

**Author notes:** Corresponding author Anirudh Mazumder, Stanford University, Stanford, California.

## Abstract

Chronic kidney disease of unknown etiology (CKDu) has emerged as an important public health challenge, particularly in agricultural communities across Southern Asia and Central America. Our research aims to explore the role of environmental factors in contributing to CKDu prevalence in these regions. Using an Extreme Gradient Boosting Machine Learning (XGBoost) model, we analyzed an environmental dataset from the CKDu endemic region of Sri Lanka. The XGBoost model achieved 85% accuracy in predicting CKDu prevalence in a total of 100 locales. Significant predictor variables included fluoride concentration in water, electrical conductivity of drinking water (EC), pH, and soil type. Soil type was the most influential factor, followed by pH and EC, which influence the solubility and bioavailability of nephrotoxic chemicals in water sources, with fluoride concentration as an additional contributing variable. The study findings highlight the need for targeted water analysis programs and interventions in water quality management, agrochemical usage, and soil treatment in CKDu-endemic regions. These insights also provide a framework for future research to identify causative agents and develop strategies for reducing CKDu prevalence.

## Introduction

Chronic kidney disease (CKD) has been extensively studied and potential target therapeutics have been developed to prevent and treat CKD in its various forms. More recently, a form of CKD, denoted CKD of unknown etiology (CKDu) has emerged [1]. Although there is incomplete knowledge of the causes of CKDu, a relationship has been shown linking CKDu and marginalized agricultural communities. In this setting the populations are affected by environmental health hazards, which are partially understood but remain largely unaddressed [1,2]. Specific factors that have been investigated in these agricultural areas include agrochemicals, heavy metals, and water source pollution [3]. These factors will drive the data exploration in the proposed project. A major affected region is in South Asia. Here we review results from analyses in South Asia and strategies identified in the literature to address this problem [4].

Machine learning (ML) techniques are a powerful approach to data analysis in many domains. ML has uniquely impacted biomedicine due to its ability to find relationships in data that are not readily identified by humans and to generate models to perform downstream tasks, including classifying different diseases based on feature sets. Using feature selectors with these ML models, one can analyze datasets to find novel biomarkers, facilitating biological understanding of disease causation [5].

Although these technologies have been used for these tasks in various contexts, they have not yet been exploited to their full potential in certain diseases, including CKDu. We sought to use the unique potential of ML models to understand specific agrochemical features correlated with CKDu, when differentiating this condition from other forms of CKD. Our goal was to develop a prediction algorithm that would predict whether a patient has CKDu or another form of CKD.

## 2. Methodology

### 2.1 Dataset

We located a dataset containing data pertaining to specific differences between CKD and CKDu patients, including environmental variables that affect these individuals, who were deidentified (Reference 6). This dataset contains data from patients in Sri Lanka, showing the unique impact that agrochemicals in that region have on the etiology of CKD.

### 2.2 Machine Learning Analysis

An Extreme Gradient Boosting (XGBoost) ML model was used for this analysis. This tool was selected for several reasons. First, the algorithm has feature selection capabilities, so the relationships the XGBoost model has learned can be interpreted using the feature selector tool, extracting the impact of various features on the dataset on outcome prediction. Second, XGBoosting algorithms can work with complex nonlinear datasets, giving these algorithms unique applicability to the data may not contain linear relationships between CKD and CKDu patients. Third, XGBoosting models are easily interpretable. This is crucial to our task, because understanding how the model makes its predictions is integral for using testing in real-world environments and to make environmental changes that may lead to reductions in CKDu case numbers worldwide.

To train the model, preprocessing steps were carried out. First, the rows from the dataset that were incomplete or where information about each feature for each subject was missing were dropped. Imputation was not used, because imputation methods could limit the real-world applicability and interpretability of the model outputs. Next, the categorical columns were encoded so they could be represented by numerical values, which is integral to the training of the XGBoost algorithm. Finally, a train/test data split was conducted, which split the training data into 80% of the dataset, and the remaining 20% were put into a testing set This approach allowed us to understand how well the XGBoost model was learning the relationship in the data. After these steps were taken, the XGBoost model was trained and feature selection was performed.

## 3. Results

### 3.1 Model Performance

In machine learning, a confusion matrix is used to evaluate the performance of classification model. Specifically, this matrix compares the values in the dataset with model-predicted values. After each iteration of the model was trained, the first metric used to assess performance of the XGBoost algorithm and to evaluate the model’s ability to understand patterns in the data was an accuracy metric. In addition to a simple accuracy metric, we used F1, Precision, and Recall metrics (**Table 1**) to quantify the model’s performance with more granularity, allowing us to change parameters, such as which class, output label, is being inspected. The F1 score integrates precision and sensitivity, to assess model performance.

**Table 1.** *Shown are* F1 scores (the harmonic mean of the precision and recall), as well as the precision and recall metrics for the XGBoost machine learning (ML) model. Precision is defined as the ratio of true positive predictions to the total number of positive predictions (true positives + false positives). Recall is defined as the ratio of true positive predictions to the total number of actual positives (true positives + false negatives).

| Class | F1 | Precision | Recall |
| --- | --- | --- | --- |
| CKD | 0.87 | 1 | 0.77 |
| CKDu | 0.82 | 0.70 | 1.00 |

The model’s accuracy, using a previously unseen test set, was 85%, indicating that the model was able to learn patterns that were influencing group assignments at a satisfactory level (**Fig. 1**). **Table 1** shows that across both classes, the model understood the relationship and how to predict that class.

**Figure 1:**
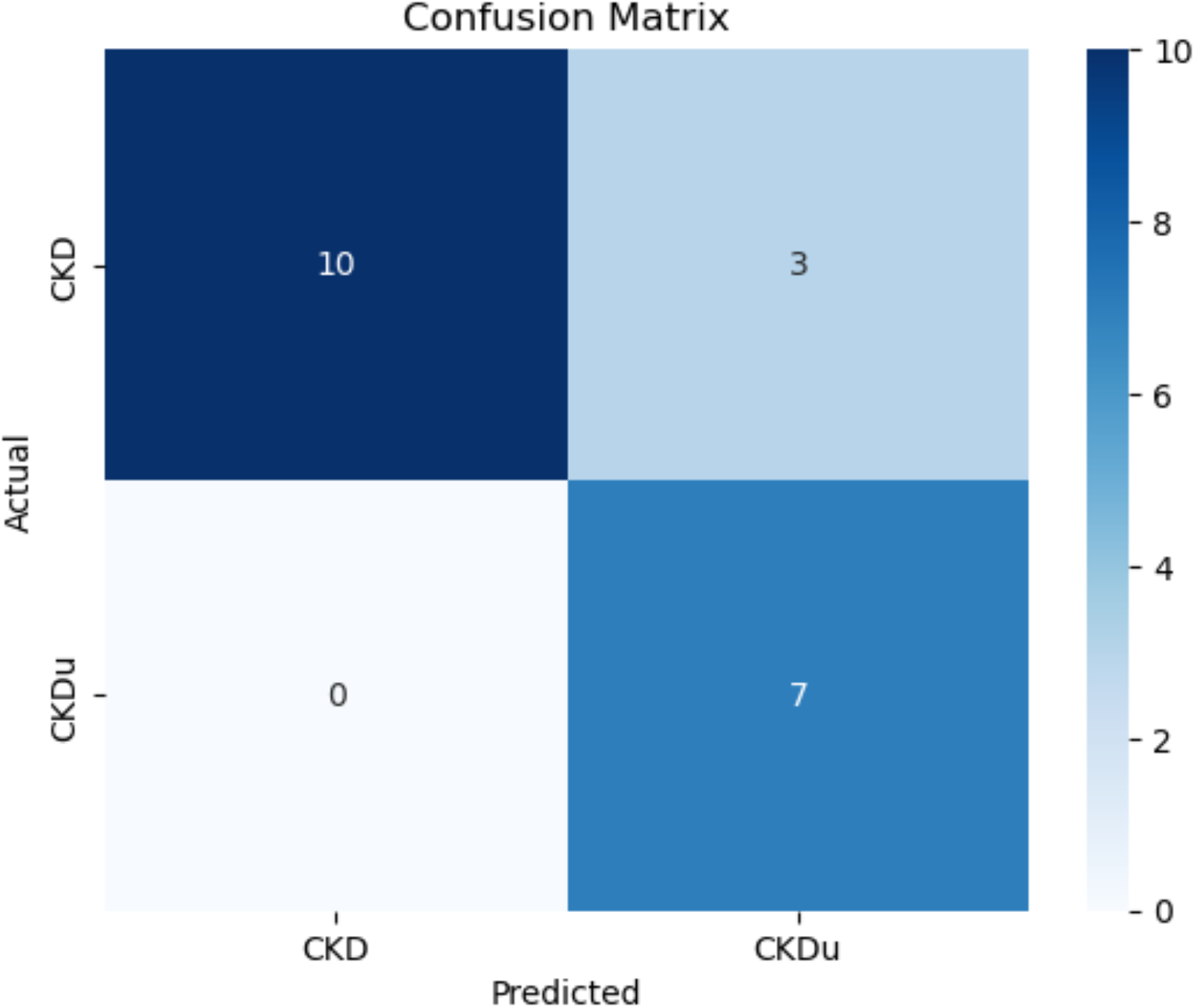
Confusion matrix showing the accuracy of the XGBoost model when making predictions on the dataset. Values from 0 to 10 refer to the number of predictions the model made in those classes.

**Figure 2:**
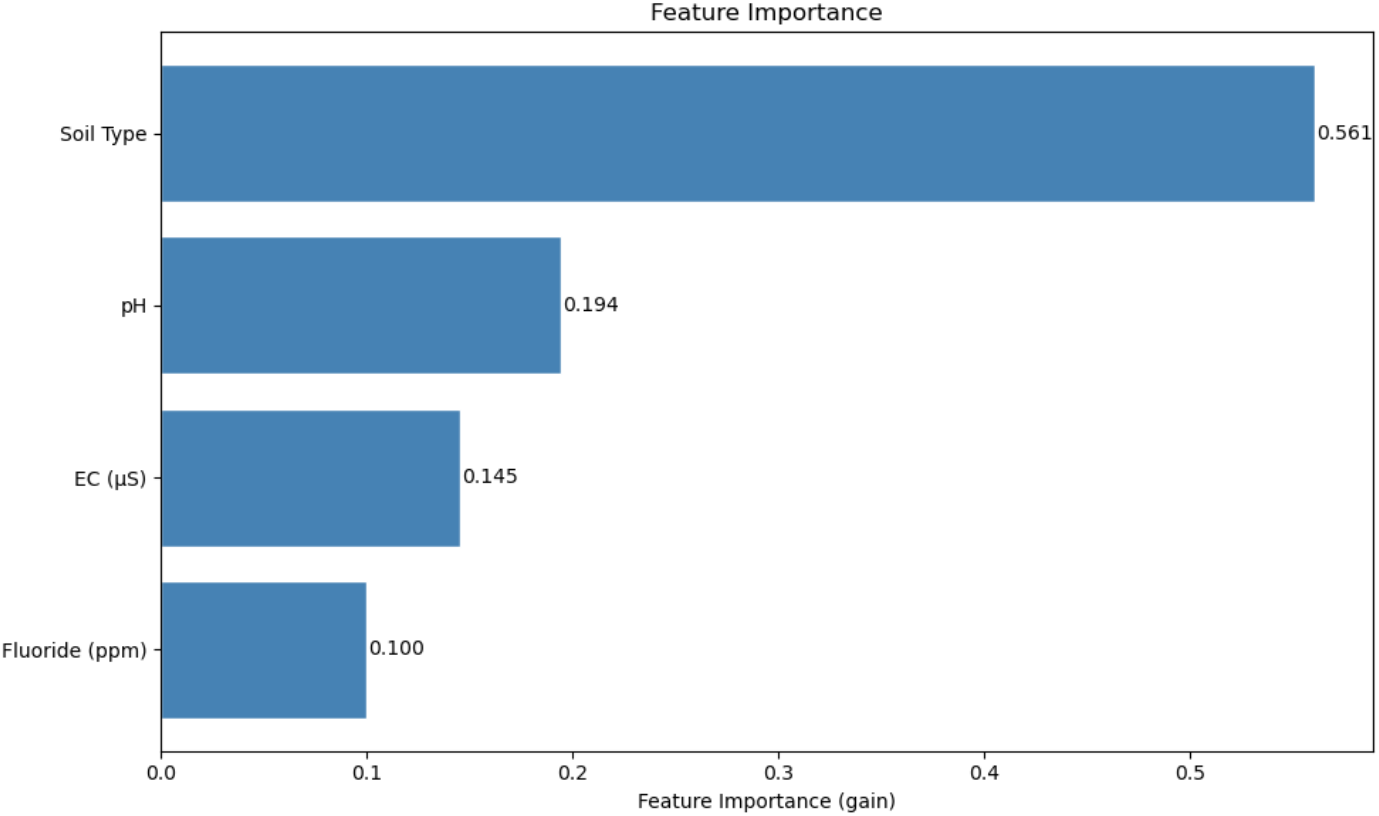
shows a graph of these values and their feature importance scores in order of model performance. This identified which features among these four variables were found to uniquely impact the outputs of the XGBoost ML algorithm, at a higher level than other features when making these predictions.

Notably, there were clear differences between the metrics for the CKD class and those for the CKDu class. The metrics indicated that the model learned how to identify CKD better than it was able to identify CKDu, which affected its predictions in the clinical dataset.

### 3.2 Feature Selection

After establishing that the ML model could learn relationships and patterns that indicate differences between CKD and CKDu, it was important to understand which features in the dataset were influential in making these predictions. The F1 score for the CKD class is 0.87, indicating a good balance between full precision (1.0) and total recall (0.77), while the CKDu class has a slightly lower F1 score of 0.82 due to lower precision (0.7) while having a better recall (1). The algorithm generated the following features as representative and potentially informative features: Soil Type, water fluoride concentration (ppm), FC (µS), pH.

## Discussion

### 4.1 Conclusion

Using an XGBoosting algorithm, we identified features in the Sri Lankan environment that correlated with differences between CKDu subjects and other CKD subjects. This algorithm will facilitate future studies by providing a framework for identifying high-risk environmental regions and particular chemicals that pose health risks.

The first and most important feature detected by the XGBoost algorithm was the soil type, followed by pH, electrical conductivity (EC), and fluoride concentration. The dataset included several distinct soil classifications, notably alluvial and reddish brown earth soils, which are characteristic of Sri Lanka’s dry zone agricultural regions. These soil types differ in their mineral composition, drainage properties, and capacity to retain agrochemicals, potentially influencing the leaching of nephrotoxic substances into local water and food sources. pH and EC, the second and third ranked features, further reflect how soil composition modulates the solubility and bioavailability of harmful minerals in drinking water. Fluoride is a common agrochemical, used in fungicides, herbicides and insecticides. It can compromise human health and may be a major cause of CKDu [7,8]. Retrospective studies have shown that fluoride is nephrotoxic, acting via diverse mechanisms. Environmental engineering solutions to filter and distill water can address this problem [9, 10]. Although the present study provides only correlations, and the findings were limited to the Sri Lankan population and further studies will be required to generalize these findings to other populations [11].

The most impactful future efforts will be to identify the specific agricultural chemicals that pollute environments in South Asia, promoting CKDu, so that these chemicals can targeted for elimination and replacement. Correlations identified by ML algorithms, as well as by other statistical techniques including those used here, cannot be assumed to be causal. Finally, as there is a high risk of confounding factors affecting the predictions of the current ML algorithm, it will be important to repeat this analysis on larger datasets with more features. Ultimately, if doubt remains about the role of these features, further data collection may be needed. Experimental work testing for toxicity maybe required. This may facilitate a deeper understanding of what agents in these environments cause CKDu.

## Data Availability

All data used by the authors is available at: https://www.kaggle.com/datasets/samanthakumara/chronic-kidney-disease-ckd-and-f

